# Acute biliary pancreatitis management during the COVID-19 pandemic

**DOI:** 10.1101/2021.05.08.21256726

**Authors:** Elif Colak, Ahmet Burak Ciftci

## Abstract

**Objective:** To analyze acute biliary pancreatitis (ABP) management during the COVID-19 pandemic.

**Methods:** This was a retrospective cohort study conducted with ABP patients during two discrete periods: a control period from March 16, 2019, through March 15, 2020 (period 1); and a COVID-19 era between March 16, 2020, and March 15, 2021 (period 2).

**Results:** A total of 89 patients with ABP were identified, 58 in period 1 and 31 in period 2, which equates to a 46.6% reduction. The mean age of the patients was 62.75±16.59 years, and 51 (57.3%) of the patients were female. qSOFA and WSES scores are significantly higher in the patients in period 2 (*p*=0.031, *p*=0.032). There were no significant differences regarding hematological parameters except lactate. Lactate levels were significantly higher in period 2 (*p*=0.012). Twenty-two patients (37.9%) in period 1 and six (19.3%) patients in period 2 underwent cholecystectomy (*p*=0.072). Cholecystectomy was performed laparoscopically in 18 (81.8%) patients in period 1 and in five (83.3%) patients in period 2 (*p*=0.932). There were no significant differences regarding surgical intervention between the two periods. Three patients were diagnosed COVID-19 in period 2. All of these patients died. The severity of ABP was significantly worse in SARS-CoV-2-positive patients, with over 100% of patients in this group developing severe pancreatitis. Six patients (10.3%) in period 1, 10 (32.2%) patients in period 2 were admitted in ICU (*p*=0.010). The median length of stay was 5 (1-40) days in period 1 and 4 (2-75) days in period 2 (*p*= 0.641). The hospital mortality rate was 3.4% and 19.3% in period l and period 2, respectively. Mortality was significantly higher in period 2 (*p*=0.012).

**Conclusion:** During the COVID-19 outbreak, a significant decrease in the number of patients with ABP and increased severity was observed. Additionally, it can be said that SARS-CoV-2 infection has a mortal course in patients with ABP. Analysis and evaluation of ABP patients during the pandemic period is important to draw conclusions that will help confront future health crises.

## Introduction

The coronavirus disease 2019 (COVID-19) pandemic has led to unexpected changes in healthcare systems across the world after the World Health Organization (WHO) declared the COVID-19 a public health emergency with a pandemic spread on March 11, 2020 [1]. In many countries, the medical care shifted according to the COVID-19 burden. Interestingly, after these changes, observed a severe decrease in non-COVID-19 patient admissions to emergency services [2-5]. Multiple studies showed a more significant reduction in emergency surgical admissions, ranging from 14% to 86% [6-9]. What happened to the emergency patients and The impacts of late or non-admissions to the hospital on emergency patients’ outcomes are critical questions.

We expected that late admission or nonadmission of patients to emergency clinics due to pandemics might lead to more complicated diseases in some cases. Studies have shown an increase in the number of complicated and perforated appendicitis during the pandemic process [10, 11]. Again, it was observed that palliative interventions other than early surgery were recommended in patients with acute cholecystitis [12, 13]. However, there has been no report investigating the effect of the pandemic on patients’ outcomes with acute biliary pancreatitis (ABP).

Acute pancreatitis is an inflammatory disease of the pancreas most commonly caused by gallstones or alcohol consumption. In most patients, the disease takes a mild course, where moderate fluid resuscitation, pain management, and nausea result in rapid clinical improvement.

Nevertheless, the severe form comprising about 20–30% of the patients is a life-threatening disease with hospital mortality rates of about 15% [14]. Initial assessment of the severity of acute pancreatitis is vital in determining further medical treatment. Intravenous fluid replacement is essential to treat fluid loss caused by third space shifts, vomiting, and increased vascular permeability caused by inflammatory mediators. Hemodynamic status should be assessed immediately upon presentation, and resuscitative measures begin as needed. Patients with organ failure should be admitted to an intensive care unit (ICU). Hydration should be provided to all patients unless cardiovascular or renal comorbidities preclude it. Early intravenous hydration is most beneficial within the first 12-24 hours and may have little benefit beyond [15-17].

In light of all these, late admission of patients with acute pancreatitis and inadequate initial treatment may increase the severity of the disease.

Thus, we aimed to investigate how a sudden disruption of seeking surgical care during the COVID-19 pandemic might affect severity rates and outcomes for ABP.

The primary aim of this study was to identify the impact of the COVID-19 pandemic on the admission and severity rates of ABP. The secondary aim was to compare the management of ABP before and during the COVID-19 pandemic.

## Methods

This single-site retrospective cohort study was conducted at the University of Samsun, Samsun Training and Research Hospital. We reviewed our hospital records of all consecutive adult patients with ABP during two discrete periods: a control period from March 16, 2019, through March 15, 2020 (period 1); and a COVID-19 era between March 16, 2020, and March 15, 2021 (period 2). The COVID-19 period was defined based on the first confirmed cases in our hospital.

Inclusion criteria included; patients over 18 years and hospital admission due to ABP diagnosed at the emergency room or during hospitalization by an imaging test (ultrasound, computerized tomography [CT]).

Exclusion criteria included; patients under 18 years, patients with acute pancreatitis without biliary etiology, patients presenting with chronic pancreatitis, pancreatic malignancy and pregnancy.

Pancreatitis was confirmed if a patient had at least two out of the following: characteristic pain, a lipase significant more than three times the upper limit of standard or radiological evidence of pancreatitis.

Disease severity was assessed with the Revised Atlanta Classification (RAC) [18]. This classification identifies two phases (early and late). Severity is classified as mild, moderate, or severe. The mild form (interstitial edematous pancreatitis) has no organ failure (OF), local or system complications and usually resolves in the first week. If there is transient (less than 48 h) OF, local complications or exacerbation of the comorbid disease, it is classified as moderate. Patients with persistent (more than 48 h) OF have a severe form of the disease. OF was defined by shock (SBP < 90 mmHg), pulmonary insufficiency (PaO2 < 60 mmHg on room air or mechanical ventilation requirement), and renal failure (serum creatinine level > 2 mg/dL or need for hemodialysis). Persistent OF was defined as lasting for over 48 hours. All acute surgical patients waiting for hospital admission and urgent surgery were screened for COVID-19 infection at the emergency department in the process of a COVID-19 outbreak in our hospital.

### Data analysis and evaluation

Records of patients with the following information were collected during the first episode of ABP on admission: age, gender, body mass index (BMI), comorbidities, Charlson comorbidity index (CCI), vital signs (body temperature, pulse rate, and systolic blood pressure [SBP]), hematology findings (white blood cell [WBC], neutrophil count, platelets, C-reactive protein [CRP], amylase, lipase, creatinine, aspartate aminotransferase (AST), alanine aminotransferase (ALT), lactate, total and conjugated bilirubin, quick sequential organ failure assessment score for sepsis (qSOFA), Bedside Index for Severity in Acute Pancreatitis score (BISAP), Glasgow-Imrie for Severity of Acute Pancreatitis, Ranson’ s criteria for pancreatitis, World Society of Emergency Surgery (WSES) Sepsis Severity Scores. In addition, the rates of magnetic resonance cholangiopancreatography (MRCP), endoscopic retrograde cholangiopancreatography (ERCP), percutaneous CT-guided fine needle aspiration biopsy (CT-guided FNAB), cholecystectomy, types of cholecystectomies, ICU admission rates, length of hospital stay, and hospital mortality were recorded.

Our study included those therapeutic interventions: endoscopic sphincterotomy, endoscopic intervention and surgical intervention. Endoscopic interventions included endoscopic drainage and necrosectomy for infected acute necrotic collection or walled-off necrosis (WON). Surgical interventions were defined by percutaneous drainage, open/laparoscopic debridement, or drainage with or without pancreatic resection. Diagnosis of WON was made according to the 2012 revised Atlanta classification; it consisted of necrotic tissue contained within an enhancing wall of reactive tissue that occurred ≥ four weeks after the onset of ABP [18].

### Outcome measures

The primary outcomes of interest were the difference in the rate of ABP admissions and severity. Secondary outcomes of interest were interventions, duration of ICU stay, length of hospital stay and mortality. The primary outcome measures were the severity of AP based on the revised Atlanta criteria. BISAP, qSOFA, Glasgow-Imrie for Severity of Acute Pancreatitis, Ranson’s criteria for pancreatitis, and WSES Sepsis Severity Scores. Secondary outcome measures included admission to ICU and length of hospital stay and hospital mortality.

### Statistical analysis

SPSS software (IBM version 20; IBM Corporation, Armonk, New York, USA) was used for statistical analysis. Scale data were tested for normality with a Shapiro–Wilk test. Non-parametric data was tested with a Mann– Whitney-test. Parametrically distributed data including were tested using a Studentt-test. Nominal data were tested using a chi-squared or Fischer exact test. A *P*-value < 0.05 was considered to indicate statistical significance.

## Results

### Patient characteristics on admission

During the study period, 89 patients with ABP were identified, 58 in period 1 and 31 in period 2, which equates to a 46.6 % reduction. The mean age of the patients was 62.75±16.59 years, and 51 (57.3%) were female. The majority (57%) of the study population was above 60 years of age. 44.9 % of patients have diabetes mellitus, 12.4 % hypertension, and 37% have no comorbid condition. 40% of the patients had BMI between 30-35. Basic demographics are outlined in Table 1. There were no significant differences in mean age (*p*= 0.767), gender (*p*= 0.372), BMI (*p*=0.181), CCI (*p*=0.538) between two periods. There were no significant differences regarding hematological parameters except lactate. Lactate levels were significantly higher in period 2 (*p*=0.012).

**Table 1.**
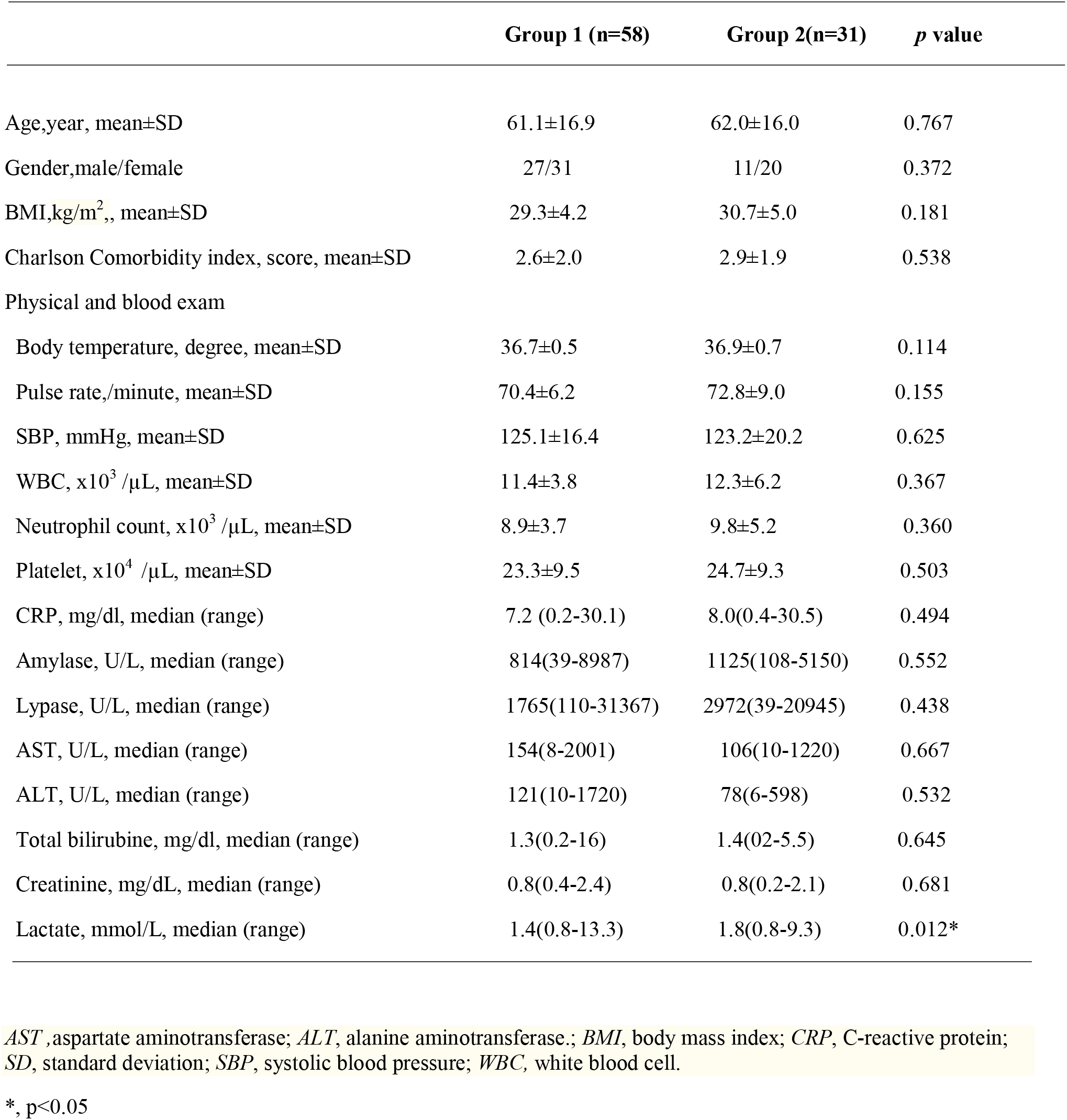
Patient demographics and baseline characteristics on admission.

|  | Group 1 (n=58) | Group 2(n=31) | *p* value |
| --- | --- | --- | --- |
| Age,year, mean±SD | 61.1±16.9 | 62.0±16.0 | 0.767 |
| Gender,male/female | 27/31 | 11/20 | 0.372 |
| BMI,kg/m<sup>2</sup>., mean±SD | 29.3±4.2 | 30.7±5.0 | 0.181 |
| Charlson Comorbidity index, score, mean±SD | 2.6±2.0 | 2.9±1.9 | 0.538 |
| Physical and blood exam |  |  |  |
| Body temperature, degree, mean±SD | 36.7±0.5 | 36.9±0.7 | 0.114 |
| Pulse rate,/minute, mean±SD | 70.4±6.2 | 72.8±9.0 | 0.155 |
| SBP, mmHg, mean±SD | 125.1±16.4 | 123.2±20.2 | 0.625 |
| WBC, x10<sup>3</sup> /μL, mean±SD | 11.4±3.8 | 12.3±6.2 | 0.367 |
| Neutrophil count, x10<sup>3</sup> /μL, mean±SD | 8.9±3.7 | 9.8±5.2 | 0.360 |
| Platelet, x10<sup>4</sup> /μL, mean±SD | 23.3±9.5 | 24.7±9.3 | 0.503 |
| CRP, mg/dl, median (range) | 7.2 (0.2-30.1) | 8.0(0.4-30.5) | 0.494 |
| Amylase, U/L, median (range) | 814(39-8987) | 1125(108-5150) | 0.552 |
| Lypase, U/L, median (range) | 1765(110-31367) | 2972(39-20945) | 0.438 |
| AST, U/L, median (range) | 154(8-2001) | 106(10-1220) | 0.667 |
| ALT, U/L, median (range) | 121(10-1720) | 78(6-598) | 0.532 |
| Total bilirubine, mg/dl, median (range) | 1.3(0.2-16) | 1.4(02-5.5) | 0.645 |
| Creatinine, mg/dL, median (range) | 0.8(0.4-2.4) | 0.8(0.2-2.1) | 0.681 |
| Lactate, mmol/L, median (range) | 1.4(0.8-13.3) | 1.8(0.8-9.3) | 0.012* |

*AST*, aspartate aminotransferase; *ALT*, alanine aminotransferase.; *BMI*, body mass index; *CRP*, C-reactive protein; *SD*, standard deviation; *SBP*, systolic blood pressure; *WBC*, white blood cell.
\*, *p*<0.05

### Severity, treatment, and clinical outcomes

The severity of index disease according to the RAC is outlined in **Table 2**. There was a significant difference between the two groups in the severity of disease in patients with ABP (*p*=0.040).

**Table 2.**
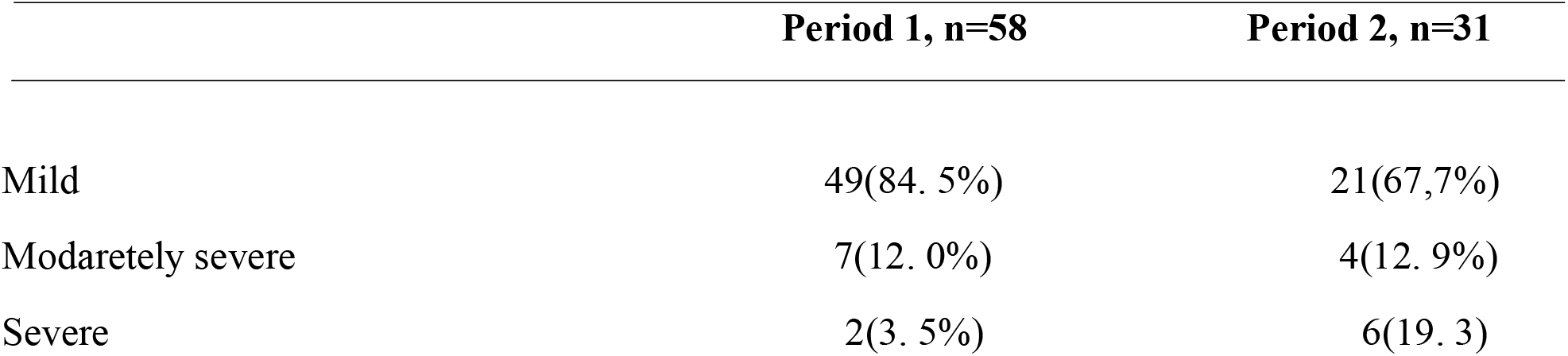
Revised Atlanta Classification of the patients in period 1 and period 2.

A comparison of the patients between two periods regarding qSOFA, BISAP, Glasgow-Imrie for Severity of Acute Pancreatitis, Ranson’ criteria for pancreatitis, WSES Sepsis Severity Scores have outlined in **Table 3**. qSOFA and WSES scores are significantly higher in the patients in period 2 (respectively, *p*=0.031, *p*=0.032)

**Table 3.** Comparison of prognostic scores between period 1 and period 2.

|  | Period 1 (n=58) | Period 2 (n=31) | *p* value |
| --- | --- | --- | --- |
| qSOFA, score, mean±SD | 0.13±0.06 | 0,38±0.13 | 0.031* |
| WSES, score, mean±SD | 0.94±0.16 | 2.03±0.44 | 0.032* |
| BISAP, score, mean±SD | 1.46±0.14 | 1.58±0.22 | 0.939 |
| Glasgow, score, mean±SD | 1.62±0.14 | 1.90±0.23 | 0,402 |
| Ranson, score, mean±SD | 2.12±0.22 | 2.90±0.34 | 0.053 |

*BISAP*; Bedside Index for Severity of Acute Pancreatitis score, *Glasgow*; Glasgow-Imrie for Severity of Acute Pancreatitis, *qSOFA*; Quick SOFA score for sepsis, *Ranson*; Ranson' criteria for pancreatitis, *WSES*; World Society of Emergency Surgery Sepsis Severity Scores.
\*, $p<0.05$

### Diagnostic and therapeutic interventions

Eighteen patients (20.2%) underwent diagnostic and therapeutic interventions that included ERCP (n=6), endoscopic drainage (n=2), percutaneous drainage (n=5) and CT-guided FNAB (n=5). Diagnostic imaging and therapeutic interventions were shown in **Table 4**. There was no significant difference found between the two periods regarding diagnostic imaging and therapeutic interventions.

**Table 4.** Comparisons of diagnostic imaging and therapeutic interventions between period 1 and period 2.

|  | Period 1 (n=58) | Period 2(n=31) | *p* value |
| --- | --- | --- | --- |
| CT | 51 (87.9%) | 30 (96.7) | 0,165 |
| MRCP | 13 (22.4%) | 10 (32.2%) | 0.312 |
| ERCP | 4 (6.8%) | 2 (6.4%) | 0.936 |
| PD | 2(3.4%) | 3 (9.6%) | 0.224 |
| CT-guide FNAB | 2 (3.4%) | 3(9.6%) | 0.224 |
| ED | 1 (1.7%) | 1 (3.2%) | 0.649 |

*CT*, Computed tomography; *CT-guided FNAB*, percutaneous CT-guided fine needle aspiration biopsy; *ED*, Endoscopic drainage; *ERCP*, Endoscopic retrograde colangiopancreaticography; *MRCP*, magnetic resonance colangiopancreaticography; *PD*, Percutaneous drainage.

Twenty-two patients (37.9%) in period one and six (19.3%) patients in period 2 underwent cholecystectomy. There was no significant difference found between the two periods regarding index cholecystectomy (*p*=0.072). Cholecystectomy was performed laparoscopically in 18 (81.8%) of cases in period 1 and in five (83.3%) cases in period 2. There was no difference found between the two groups regarding laparoscopic cholecystectomy (*p*=0.932).

### Outcomes

Six patients (10.3%) in period 1, 10 (32.2%) patients in period 2 were admitted in ICU. ICU admission rates were significantly higher in period 2 (*p*=0.010). The median length of stay was 5 (1-40) days in period 1 and 4 (2-75) days in period 2. There was no difference found between the two groups (p= 0.641). Two patients in period 2 were diagnosed COVID-19 at admission; one patient was diagnosed COVID-19 after admission (7th day). All of these patients died. The severity of ABP was significantly worse in SARS-CoV-2-positive patients, with over 100% of patients in this group developing severe pancreatitis. The hospital mortality rate was 3.4% and 19.3% in period l and period 2, respectively. Mortality was significantly higher in period 2 (*p*=0.012).

## Discussion

ABP is the most common form of acute pancreatitis encountered by physicians in emergency departments globally. This study has shown that significantly fewer patients with ABP were admitted during the COVID-19 pandemic than the pre-pandemic period. The findings of our study are similar to those seen in the UK, Italian, and Spanish studies, which found significant decreases in general surgical admissions, ranging from 14% to 86% [4-6]. Swagman et al. also reported a significant decrease in medical emergencies in all disciplines since introducing contact restrictions during the COVID-19 pandemic, according to their analysis of data from 36 emergency services [2].

Nevertheless, this could be related to the public measures implemented to reduce mobility, the hesitation of going to emergency services due to contagion risk, and the spontaneous resolution of mild ABP self-limited with symptomatic treatment at home. It may reduce the overall ABP diagnosis, and it might increase the severity of ABP presentation.

Second, we expected patients to present with a delayed and potentially more acute during the COVID-19 pandemic. Because initial treatment is vital in managing ABP, aggressive intravenous fluid replacement should treat fluid loss caused by third space shifts, vomiting, and increased vascular permeability. We found that lactate levels are significantly higher in patients in period 2 on admission. It could be related to inadequate fluid replacement.

Morever, prompt evaluation of the severity of ABP is essential to predict patients’ outcomes, estimate prognosis, and determine the need for ICU care.

In addition, we found that qSOFA and WSES scores are higher for patients in period 2 than period 1. Clinical signs with higher qSOFA and WSES scores indicate a higher rate of more severe disease courses concerning the whole included patient population. A similar situation has been demonstrated for patients with acute appendicitis. Many studies have shown a decrease in acute appendicitis admissions as well as an increase in the number of complicated appendicitis during the COVID-19 pandemic [10, 11]. Willms et al. reported that the overall number of patients decreased from 1027 in 2019 to 888 (−13.5%, p = 0.003), and the rate of complicated appendicitis rose to 64.4% (p=0.012) during the lockdown [11].

In the present study, we found that three of the six patients who died in period 2 had concurrent COVID-19 infection. All three patients had persistent OF and necrotizing pancreatitis. It is difficult to make a definitive interpretation of the cause of death, as both diseases can cause OF. However, we can conclude that the coexistence of COVID-19 and ABP can lead to severe illness and death.

This study also describes the management of ABP before and during the COVID-19 pandemic. Although the number of our patients was small, we found that we performed cholecystectomy in both periods at a similar rate in patients with mild ABP. Cholecystectomy was performed laparoscopically in 18 cases in period 1 and 5 cases in period 2. No change to surgical management of mild ABP was implemented in our clinic in response to the possible increased risk of exposure to COVID-19 from laparoscopic surgery. All of these patients were discharged in good condition.

We also used advanced imaging modalities and interventional procedures such as MRCP, ERCP, CT-guided FNAB, percutaneous drainage, endoscopic drainage at similar rates in both periods. It can be interpreted as the standard of emergency surgical care for patients with ABP was maintained in our hospital.

The main limitation is that it is a retrospective study, where the electronic medical record was obtained retrospective data, and the interpretation of these data might suppose a bias. In addition, it has been influenced by interindividual variability in clinical decision-making.

Despite many surgical societies’ recommendations about ABP management, there is no clear evidence about ABP management during the COVID-19 pandemic. In this paper, we describe and analyze ABP management in our center during the higher incidence period of the pandemic to draw conclusions that help to face future health crises in emergency surgical patient management.

To our knowledge, this is the first study that analyzed the number, severity, and management of ABP during the COVID-19 pandemic.

## Conclusions

There was a significant decrease in patients with ABP applied to our hospital during the COVID-19 pandemic. In the other hand, there was an increase in the severity of the disease was observed. This increase in the severity of the disease may be that the initial treatment did not commence at the appropriate time. In addition, it can be said that SARS-CoV-2 infection has a mortal course in patients with ABP. Analysis and evaluation of ABP patients during the pandemic period is important to draw conclusions that will help confront future health crises. Thus, multicenter studies with more significant numbers of patients are needed to obtain more valuable evidence regarding ABP management during the COVID-19 pandemic.

## Data Availability

Data available from author on request.

## Ethical approval

This study was approved by the Review Board of the University of Samsun, Samsun Training and Research Hospital.

